# Longitudinal Transdisciplinary Neuropalliative care Support (LOTUS) Study – a conceptual framework and fidelity assessments

**DOI:** 10.64898/2026.05.29.26354486

**Authors:** Claire J. Creutzfeldt, Ann Leonhardt-Caprio, Elizabeth Nielsen, Robert Y. Lee, Sarah Wahlster, Robert G. Holloway, Lynn F. Reinke

## Abstract

**Importance:** Severe stroke is a leading cause of death and disability worldwide. Survivors and their families face long-term unmet needs, including care that does not reflect patients’ values, fragmented care, and high rates of psychological distress among caregivers.

**Objective:** To describe the conceptual framework of the longitudinal transdisciplinary neuropalliative care support (LOTUS) intervention and assess its fidelity in a pilot feasibility study.

**Design:** Pilot feasibility randomized study; fidelity was assessed using weekly checklists completed by the LOTUS nurse and qualitative analysis of weekly LOTUS team meeting transcripts.

**Setting:** Single comprehensive stroke center in Western New York.

**Participants:** Patients hospitalized with severe stroke and their caregivers. Dyads were randomized to usual care or intervention.

**Intervention:** The LOTUS intervention is implemented in a stepped-care fashion using 5 strategies: Awareness, Assistance, Adjustment, Acceptance and Alignment (5A’s). Led by a specially trained nurse with a chaplain, social worker, psychologist, and neuropalliative care physician, the LOTUS team follows dyads from early in the hospital course through 6 months.

**Main Outcomes and Measures:** Fidelity, the degree to which the intervention was delivered as intended, assessed via (1) utilization of 5A activities from weekly LOTUS checklists; (2) thematic analysis of weekly LOTUS team meeting transcripts.

**Results:** Of 26 patients in the trial, 13 were randomized to intervention. The LOTUS nurse completed 108 checklists, with an average of 619 minutes of direct contact per participant over 6 months. Each component of the 5A’s was utilized. Awareness and Assistance predominated early after enrollment and revolved around personhood, support, and self-efficacy. Adjustment was especially relevant during care transitions and was typically supported by the LOTUS social worker. Acceptance and Alignment were more prevalent during later meetings, with the LOTUS psychologist supporting identification and modeling of coping skills and the LOTUS physician guiding prognosis and goals-of-care conversations. The LOTUS nurse served as primary point of contact, providing continuity and a trusting relationship, while other team members functioned in a predominantly advisory role.

**Conclusions:** The LOTUS intervention was delivered with fidelity to the 5A-framework, supporting a future randomized clinical trial to evaluate its efficacy in patients with severe stroke and their caregivers.

## Introduction

Severe stroke accounts for over 6 million deaths annually and is a leading cause of functional and cognitive disability worldwide.^1^ Patients often present with profound neurological impairments, while their families are confronted with sudden and significant loss, compounded by prognostic uncertainty and the burden of making treatment decisions that align with the patient’s values. While often considered an acute event, severe stroke is a chronic condition where survivors and their family are faced with long-term challenges and unique, unmet palliative care needs including steep declines in quality of life, receipt of care that does not reflect patients’ values and preferences, and high rates of anxiety and depression among family caregivers that persist for many months following hospitalization.^2–4^ Untreated caregiver burden and depression are associated with lower quality of care and worse outcomes for both caregivers and survivors, and guidelines recommend that interdisciplinary team members address and support caregivers and patient-family dyads.^5–7^

Palliative care has developed both as a distinct medical specialty and a core clinical skillset, aiming to enhance communication about goals-of-care and promote quality of life for patients and their families. For patients with cancer, palliative care interventions have been shown to improve symptom burden and quality of life and improve caregiver outcomes,^8^ and this benefit may, in part, be mediated by increased use of effective coping skills.^9,10^

Based on the social care model described by the National Academies of Science and our own work,^2,11–13^ we developed a novel intervention specific to severe stroke, the longitudinal transdisciplinary neuropalliative care support (LOTUS) intervention. This model emphasizes the need to provide patient- and family-centered longitudinal care; to adjust care delivery to an individual’s biopsychosocial context; and to develop effective transdisciplinary teams and communication pathways across care settings.^11^ We conducted a pilot feasibility study of the LOTUS intervention to establish proof of concept and assess implementation.

This article describes (1) the conceptual framework that underpins this innovative model and its proposed mechanisms of action. We then present (2) fidelity findings from our pilot study — that is, the extent to which the intervention was delivered as intended^14^ — to inform refinements in preparation for a future efficacy trial and potential clinical adoption within healthcare settings.

## Methods

Patients were eligible if they had a severe stroke as the primary cause of hospital admission (ischemic stroke, non-traumatic intracerebral hemorrhage or subarachnoid hemorrhage), defined as Glasgow coma scale ≤ 12 or NIH Stroke scale ≥10 for ≥ 48 consecutive hours in the first 7 days of hospitalization. Caregivers, defined as family or friends, were eligible if they were ≥ 18 years, English-speaking, and self-identified as involved in the unpaid care of the patient. Patient-caregiver dyads were randomized (in variable-sized blocks using a computer-based algorithm) to usual care or intervention. Subjects assigned to intervention worked with the LOTUS team and completed outcome assessments, subjects assigned to control completed outcome assessments only. The University of Rochester institutional review board approved this study (STUDY00009703).

### Description of the LOTUS intervention

The LOTUS intervention is implemented in a stepped-care fashion using 5 strategies: Awareness, Assistance, Adjustment, Acceptance and Alignment (5A’s, Table 1). Led by a specially trained nurse with other team members including a chaplain, a social worker, a psychologist, and a neuropalliative care physician, the LOTUS team follows patients with severe stroke and their caregivers from early in their hospital course through 6 months post-enrollment with the main objectives to (1) promote self-efficacy and coping skills for caregivers facing severe stroke; (2) ensure treatment is aligned with patients’ goals-of-care throughout their illness trajectory; and (3) improve patient and caregiver long-term outcomes. The LOTUS nurse serves as primary point of contact and meets with participants daily during the intensive care unit (ICU) stay, every other day during acute care, and weekly after hospital discharge, including in rehabilitation facilities, nursing facilities or at home. Following hospital discharge, meetings may be in person or virtual (Zoom/telephone). The nurses’ actions are guided by a *checklist* containing activities from each of the 5A’s that is completed weekly (supplement). The nurse documents the amount of direct contact with participants and the content focus of these contacts. Each of the other LOTUS team members dedicates on average 2 hours per week to LOTUS activities. One hour is allocated for individual tasks such as collaborating with or supporting the LOTUS nurse, or direct interaction with participants. The other is reserved for a *weekly virtual LOTUS team meeting* focused on case review. This weekly meeting brings the nurse and their transdisciplinary team together to discuss each participant’s needs, challenges, and progress in detail. The nurse outlines actions taken, highlights areas where they are encountering difficulties, and identifies situations requiring additional guidance. Using the 5A framework, team members offer structured support and expert consultation to refine care strategies, strengthen clinical decision-making and discuss next steps.

**Table 1.** 5 A’s conceptual framework for the LOTUS intervention and specific goals for each “A”. *SuPPOrTT questions: Su: Does the patient/family need psychosocial or spiritual <u>S</u>upport or help with coping? P: Does the patient have <u>P</u>ain or other distressing symptoms? PO: Do patient/family have questions about <u>P</u>rognosis or treatment <u>O</u>ptions? rTT: Do we need to <u>r</u>eaddress goals of care or <u>T</u>arget <u>T</u>reatment to patient-centered goals?^2^ **Self-efficacy: based on Bandura’s Social Cognitive Theory, self-efficacy is a person’s own judgment of their ability to succeed at a specific task or achieve a desired outcome, in this context caregiving or getting questions answered from the healthcare team.^23^ ***Radical acceptance: is a core skill in dialectical behavioral therapy that helps one fully accept one’s reality and stop fighting unchangeable facts so that one can respond more effectively and constructively and enable change.^24^ ****Dialectical Behavioral Therapy: psychological therapy that teaches skills to help deal with intense emotions with a focus on balancing acceptance and change.^25^

|  |  |
| --- | --- |
| **Awareness** | Identify evolving stroke-specific patient/caregiver needs using the SuPPOrTT questions* with a goal of building trust and understanding, recognize the trajectory and explore how we can best assist. |
| **Assistance** | First response to identified SuPPOrTT* needs, i.e. the LOTUS RN provides practical care and coping skills, resource navigation and coordination, explores goals-of-care and looks for opportunities for a time-limited trial. LOTUS RN models and teaches self-efficacy** |
| **Adjustment** | Care coordination, communication & decision support <u>across care facilities</u>. The LOTUS team helps the caregiver utilize the healthcare system to meet the specific needs of the participant with a particular focus on times of transitions. Patients and families learn new roles and routines. |
| **Acceptance** | Support and teach advanced coping skills using principles of Dialectical Behavioral Therapy (DBT); helping families cope with uncertainty, grief, loss; meaning-making. Acceptance involves embracing reality and the associated emotions as they are, using evidence-based strategies like mindfulness, DBT skills, adaptive coping techniques and ultimately 'radical acceptance'*** |
| **Alignment** | Ensure 'everyone is on the same page' (dyad-team unity). This means ensuring prognosis concordance (agreement between all individuals about the patient's expected outcome or range of outcomes) and goal concordance (agreement between a treatment goal and a person's core values, interests and identity). LOTUS explores prognostic understanding and goals-of-care iteratively. A time-limited trial is re-visited, advance care planning is encouraged. |

For this study, the LOTUS nurse was an RN with >10 years of nursing experience across primary, chronic and neuro-ICU care including stroke. For the role as LOTUS nurse, training included self-learning modules, online (VitalTalk) and in-person workshops covering the LOTUS protocol, stroke-specific palliative care, and collaborative care (supplement).

This pilot study was conducted at a single comprehensive stroke center in Western New York. The LOTUS nurse, chaplain and social worker were local to New York State, the neuropalliative care physician and psychologist were at a different site.

#### Fidelity

Fidelity in intervention research refers to the degree to which an intervention was delivered as intended and is typically assessed through observational field notes, logs and recordings of sessions.^15^ We used checklists completed by the LOTUS nurse and recordings of the LOTUS team meetings.

a. Checklists (supplement). From the weekly checklists, we extracted the duration of direct contact with participants per week as well as utilization of 5A-activities during these contacts and the LOTUS RN’s rating of participant engagement from 1 (little to none) to 5 (great deal).
b. Weekly virtual LOTUS team meetings were recorded and transcribed automatically to assess implementation of the “5A’s” framework and provide in-depth evaluations of the challenges experienced by caregivers of patients with severe stroke over time, how the LOTUS nurse responded to those, how the LOTUS nurse felt they were able to meet family needs and where they needed advice from the LOTUS Team. From these transcriptions, we created one document per individual case that included all LOTUS conversations relevant to that participant. These transcripts were reviewed by our analytic team of two neurologists, a nurse investigator, and a neuroscience student, bringing diverse clinical and research perspectives to the interpretive process and enhancing the depth and rigor of the analysis. Each investigator read at least 8 of the 13 transcripts with each transcript read by at least 2 investigators. We employed a hybrid coding strategy: deductive coding guided by the 5A’s framework to structure our initial analysis, inductive coding to identify additional emergent themes beyond the framework. Deductive coding served as a “top-down” approach by using the 5A’s as pre-determined codes and then applying those to the data to test, confirm, or refine the theory. This approach focuses the analysis on finding evidence for or against the existing framework.^16^

Transcripts were reviewed collaboratively, with regular meetings to discuss coding decisions, resolve discrepancies, and refine thematic categories through iterative consensus-building.

## Results

Of the 26 patients included in the trial, 13 were randomized to intervention (table 2) and described here as the purpose of this analysis was to establish fidelity of the intervention.

**Table 2.** Intervention Participant Characteristics.

|  | Patient<br>N=13 | Caregiver<br>N=13 |
| --- | --- | --- |
| Age, mean (SD), years | 67.2 (15.7) | 51.75 (16.3) |
| Female participants, n (%) | 8 (61.5%) | 10 (76.9%) |
| Race, n (%) |  |  |
| - Asian | 1 (7.7%) | 1 (7.7%) |
| - Black | 2 (15.4%) | 2 (15.4%) |
| - White | 10 (76.9%) | 8 (61.5%) |
| - Other | 0 (0%) | 0 (0%) |
| - Missing | 0 (0%) | 2 (15.4%) |
| Ethnicity, n (%) |  |  |
| - Hispanic | 0 (0%) | 0 (0%) |
| - Non-Hispanic | 13 (100%) | 13 (100%) |
| - Missing | 0 (0%) | 0 (0%) |
| Diagnosis, n (%) |  |  |
| - Ischemic stroke | 10 (76.9%) |  |
| - Intraparenchymal hemorrhage | 2 (15.4%) |  |
| - Subarachnoid hemorrhage | 1 (7.7%) |  |
| NIHSS on admission, median (range) | 20.5 (15-30) |  |
| Follow-up NIHSS (between day 2 and randomization), median (range) | 16 (11-27) |  |
| Total length of stay in hospital, days, median (range) | 21 (7-52) |  |
| mRS at hospital discharge, median (range) | 4 (4-6) |  |
| Discharge location, n (%) |  |  |
| - Home | 1 (7.7%) |  |
| - IPR | 5 (38.5%) |  |
| - SNF (= subacute Rehab or long-term care)<br>- Deceased in hospital | 4 (30.7%)<br>3 (23.1%) |  |
| Residence at end of intervention (6 months), n (%)<br>- Home<br>- "In a rehab facility"<br>- "In a nursing home for long term care"<br>- Deceased<br>- Other | 5 (38.5%)<br>2 (15.4%)<br>1 (7.7%)<br>4 (30.7%)<br>1 (7.7%) |  |
| Relationship to patient, n (%)<br>- Spouse/partner<br>- Parent<br>- Adult Child<br>- Other<br>- Missing/Unanswered |  | 6 (46.2%)<br>1 (7.7%)<br>3 (23.1%)<br>2 (15.4%)<br>1 (7.7%) |
| Lived with patient prior to SABl (n, %) |  | 7 (53.8%)<br>{unanswered = 1} |

### Checklists

The LOTUS nurse completed 108 checklists in total suggesting an average of 619 minutes of direct contact per participant over 6 months (supplemental table 3).

### Meeting transcripts

Because Awareness encompasses the identification of needs and Assistance is the immediate (basic) response to Awareness, we combined these two in our report. While Awareness and Assistance tended to take up more space early after participant enrollment and were mainly represented by the LOTUS nurse, Adjustment, Acceptance and Alignment were more prevalent during later meetings and offered more dialogue with LOTUS team members.

#### Awareness/assistance

As the LOTUS nurse got to know and build relationship with participants, weekly reports back to the LOTUS team revolved around 3 main themes: personhood, support and self-efficacy (table 4). *Personhood* encompasses the stories that illustrate who the patient is, who the caregiver is and how they relate to each other and their environment, participant’s support system and past experiences, be they trauma, tragedy, mental or physical health issues. The LOTUS nurse used a strength-based approach by recognizing caregivers’ abilities to manage previous challenging experiences and apply those skills to the current situation *(“You have this horrible thing that happened* {*domestic abuse*}, *but it’s prepared you to have all of these amazing skill sets”*). Stories evolved over time, as relationships with caregivers deepened and situations changed. *Support* needs were varied, and the LOTUS nurse could tailor their support to the individual’s needs. Examples of *Self-efficacy* evolved mostly around encouraging the caregiver to speak up to the medical team and make their voices heard.

**Table 4.**
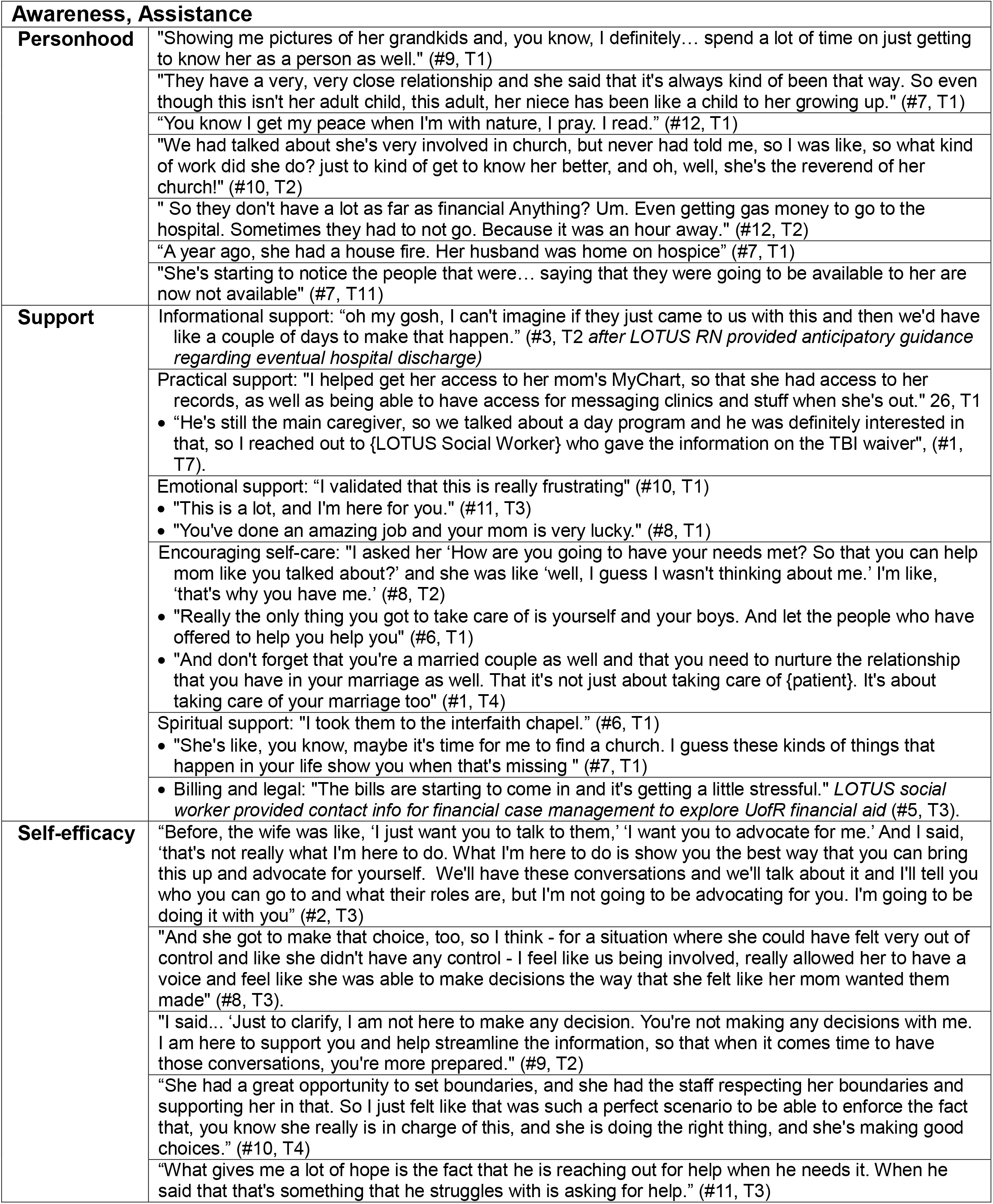
Example quotes for the first two A’s: Awareness and Assistance are the basic identification and response to patient or family needs. ‘T’ is the timepoint for this quote, e.g. T4 would be the fourth Monday that this participant was discussed.

#### Adjustment

Early adjustment after the acute phase often involved navigating discharge to a nursing or rehabilitation facility. This transition required assistance with care coordination, communication and decision support across facilities, including the realization that this transition was the appropriate next step. Knowing how the system works, the LOTUS nurse, with support from the LOTUS social worker, could make things happen that may not have happened otherwise: *“I’ve been watching her therapy notes because she initially didn’t qualify for inpatient rehabilitation (IPR). … I messaged the nurse practitioner and said: ‘What do you think about reassessing her since she hasn’t gotten a bed offer?’ … and I see the discharge order (to IPR) just went in this morning!”*.

Another patient had decided that her time-limited trial after her stroke was completed and was refusing food and drink at home. Given several previous conversations, the LOTUS team discussed how consistent this was with her overall values and her messaging since the stroke which had left her with severe aphasia. *“LOTUS MD: I’m going to talk with* {*Hospice*} *today about setting something up” - Chaplain: “I will*…*meet with her and/or her daughter*.*”*

#### Acceptance

Caregivers exhibited and learned various coping strategies (table 5). Several observations highlighted that acceptance can happen only when one allows space for emotions: *Psychologist: “some of these moments where* {*participant*} *have been able to express emotion have been happening more and that’s probably the thing that needs to happen in order for them to move closer towards acceptance - You’re doing a great job of keeping that in mind and* .. *validating that. I would imagine that allowing that grief to fully be there is probably what’s going to allow them to move towards a place of acceptance for the first time*.*”* In the subsequent visit, the LOTUS psychologist helped the LOTUS nurse address the participant’s tendency to avoid emotion: *“first, see if you describe your observation in very nonjudgmental language, would she agree with it: ‘I noticed that when certain topics are brought up, you express an unwillingness, or lack of interest in talking about those things’, and see if they will agree, and then say, ‘I was wondering if we could talk about that and how is that helping you right now? And how is that important to your coping? Are there any ways in which you feel like not being willing to talk about those things is creating more challenges in this situation?”*

**Table 5.**
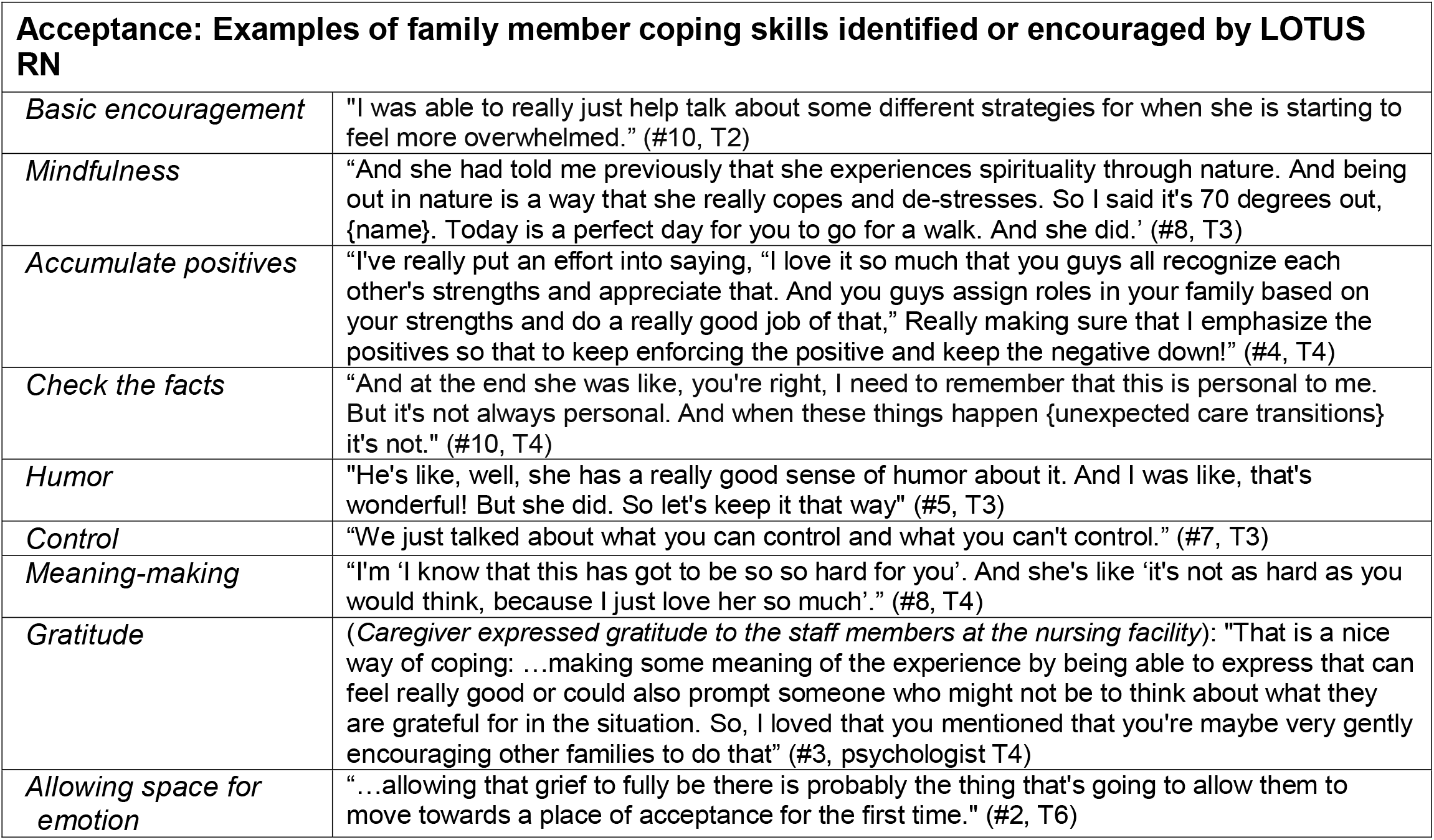
Example coping strategies and quotes for Acceptance as stated by LOTUS RN in Monday meetings. ‘T’ is the timepoint for this quote, e.g. T4 would be the fourth Monday that this participant was discussed.

| **Acceptance: Examples of family member coping skills identified or encouraged by LOTUS RN** |  |
| --- | --- |
| *Basic encouragement* | "I was able to really just help talk about some different strategies for when she is starting to feel more overwhelmed." (#10, T2) |
| *Mindfulness* | "And she had told me previously that she experiences spirituality through nature. And being out in nature is a way that she really copes and de-stresses. So I said it's 70 degrees out, {name}. Today is a perfect day for you to go for a walk. And she did.' (#8, T3) |
| *Accumulate positives* | "I've really put an effort into saying, "I love it so much that you guys all recognize each other's strengths and appreciate that. And you guys assign roles in your family based on your strengths and do a really good job of that," Really making sure that I emphasize the positives so that to keep enforcing the positive and keep the negative down!" (#4, T4) |
| *Check the facts* | "And at the end she was like, you're right, I need to remember that this is personal to me. But it's not always personal. And when these things happen {unexpected care transitions} it's not." (#10, T4) |
| *Humor* | "He's like, well, she has a really good sense of humor about it. And I was like, that's wonderful! But she did. So let's keep it that way" (#5, T3) |
| *Control* | "We just talked about what you can control and what you can't control." (#7, T3) |
| *Meaning-making* | "I'm 'I know that this has got to be so so hard for you'. And she's like 'it's not as hard as you would think, because I just love her so much'." (#8, T4) |
| *Gratitude* | (*Caregiver expressed gratitude to the staff members at the nursing facility*): "That is a nice way of coping: ...making some meaning of the experience by being able to express that can feel really good or could also prompt someone who might not be to think about what they are grateful for in the situation. So, I loved that you mentioned that you're maybe very gently encouraging other families to do that" (#3, psychologist T4) |
| *Allowing space for emotion* | "...allowing that grief to fully be there is probably the thing that's going to allow them to move towards a place of acceptance for the first time." (#2, T6) |

A barrier to acceptance is ambiguity, which participants experienced in multiple ways, typically due to prognostic uncertainty. One family member expressed distress in their role as surrogate decision-maker whose mother’s range of possible outcomes was wide. LOTUS nurse paraphrased their feelings as *“I’m fearful I make the wrong decision and she dies and it’s not what she would have wanted*.*”* Another struggled with the recovery journey her father was enduring: *“I don’t want to let my dad die, but I also want my dad to die*.*”* Finally, families also felt role ambiguity in the face of new and conflicting social roles or identities with unclear boundaries.

#### Alignment

The LOTUS nurse encouraged goals-of-care conversations and educated families about goal concordance. Similar to acceptance, the LOTUS nurse recognized that, especially early in the hospital course and in the setting of prognostic uncertainty, Alignment had to be a future goal to achieve rather than an immediate need to be met:

*“The patient* .. *didn’t want to live if she had to depend on anybody. She had told her daughter that she wanted her to withhold food and hydration, if that were to happen. And now* {*daughter*} *is like ‘I just don’t know what to do because right now I feel like I want her to go to rehab and get strong so she can come home and have some quality of life, but I don’t know if that’s really consistent with what mom* {*wants*}.*”*

While some needed the patient’s prior voice to reach alignment (as above), others preferred to wait for the patient’s ‘future self’ to express their preferences: *“I didn’t get a feeling that they even want the voice from the patient from 6 months ago. That doesn’t seem to be leading them in their decision-making*… {*Participant*} *was very clear about that – ‘you know, who knows what he’s going to be like, and nobody can judge that’”(MD)*.

The LOTUS MD commonly advised on how to approach goals-of-care conversations: *“I like to twist that question a little bit by making sure people understand that it is asked usually as a treatment preference. Do you want this treatment or not? But what it should be is what we would call an outcome preference: Are you willing to undergo this treatment if the outcome is going to be X? After stroke, it is always a range of outcomes. And so, the way most people make these decisions is that if that range, even the best-case scenario does not seem acceptable to her, then the PEG tube does not seem like a good idea”*. The LOTUS MD also assisted the LOTUS nurse in providing anticipatory guidance and encouraged revisiting goals-of-care conversations in the post-acute setting.

#### Other observations

Other observations from the meeting transcripts review evolved around the role of the LOTUS team members as well as the LOTUS nurse’s insight into her own role, including a learning curve over time: *RN: “One of the things I struggle with is, I tend to go into problem-solving mode…” MD: “I think one beautiful word is curiosity. To allow curiosity, too*.*” Psychologist: “I like that idea - instead of your goal being to take away their pain, it’s to understand what they’re feeling - ask open-ended questions …” RN: “It’s definitely part of the learning curve for me, is switching that mindset of being a fixer*.*”*

While the breadth of skills necessary to manage participant’s diverse needs required the complementary expertise of the LOTUS team members, their role was more advisory to the LOTUS nurse with only rare direct interactions with participants. That responsibility fell to the LOTUS nurse, the primary point of contact, who provided continuity and built a supportive, trusting relationship.

## Discussion

After severe stroke, caregivers encounter complex challenges that necessitate responsive, individualized support guided by a structured framework and adaptable to changing needs over time. The 5As provided a transparent and standardizable roadmap for the LOTUS nurse in their regular encounters, as well as for the LOTUS team in their weekly meetings. Our qualitative analysis suggests that each of the components addressed important challenges and each was utilized, meeting fidelity requirements. This transdisciplinary care model has demonstrated effectiveness in other palliative care populations including advanced cancers and chronic diseases, supporting the adaptation to people faced with severe stroke.^17–19^

While the LOTUS intervention was designed as a “stepped care” model, the 5A’s were applied fluidly, depending on the participant’s situation. Awareness and Assistance were present in most encounters, whereas Adjustment was especially relevant during care transitions and was typically supported by the LOTUS social worker. The LOTUS psychologist supported and helped the LOTUS nurse identify and encourage different coping skills for different people and as coping needs evolved across the illness trajectory. Effective coping preserves psychological integrity,^10^ making coping-skills modeling central to this intervention. Aiming for prognosis concordance (shared understanding of likely outcomes^20^) and goal concordance (care aligned with patient values^21,22^), alignment cannot be achieved through a single conversation; it is an aim pursued across the illness trajectory, during which patients, their families, and clinicians come to understand the patient’s core values, witness the gradual adaptation that serious illness demands,^13^ and, ideally through longitudinal transdisciplinary support, find peace and acceptance with how those values align with treatment options and outcomes.

The weekly LOTUS team meetings were central to leveraging the team members’ expertise. While the study was designed to give each member time to meet with individual participants, the nurse’s role as liaison between participants and the expert team appeared preferred. A hybrid version, where the nurse brings a LOTUS team member with her, may offer the optimal balance of relational continuity and specialist input.

This fidelity presentation needs to be seen in the context of limitations, including the single site and small sample size that preclude generalizability, and fidelity reflects only one stage of implementation and does not indicate acceptability, feasibility or sustainability. Two additional articles from this group supporting acceptability and feasibilty are in process.

Highlighting the conceptual model of the LOTUS intervention and the results of fidelity assessments, this article presents a highly innovative example of providing transdisciplinary support to patients with severe stroke and their families using a theory-driven foundation to support self-efficacy^23^ and a framework to make the multifaceted and complex intervention replicable. This study supports a future randomized clinical trial to evaluate the efficacy of the LOTUS intervention. Scaling these analyses to a RCT will require natural language processing of session and team-meeting transcripts, which could additionally characterize the timing of each A, the populations served, and explore associations between delivery patterns and outcomes.

## Data Availability

All data produced in the present study are available upon reasonable request to the authors

## Acknowledgements

We would like to thank Melissa Christodaro, Mira Reichman, Rachel Christopher and Kelly Spahr for their invaluable and compassionate expertise as LOTUS team members, and student Misako Wongpa for her assistance with the transcripts and early analysis.

## References

1. Martin SS, Aday AW, Allen NB, Almarzooq ZI, Anderson CAM, Arora P, Avery CL, Baker-Smith CM, Bansal N, Beaton AZ, et al. 2025 Heart Disease and Stroke Statistics: A Report of US and Global Data From the American Heart Association. Circulation. 2025;151:e41–e660. doi: 10.1161/CIR.0000000000001303

2. Plinke WV, Buchbinder SA, Brumback LC, Longstreth WT, Kiker WA, Holloway RG, Engelberg RA, Curtis JR, Creutzfeldt CJ. Identification of Palliative Care Needs and Mental Health Outcomes Among Family Members of Patients With Severe Acute Brain Injury. JAMA Netw Open. 2023;6:e239949. doi: 10.1001/jamanetworkopen.2023.9949

3. Smith NL, James A, Matin N, Fong CT, Sharma M, Lele AV, Robba C, Mazwi N, Wiseman DB, Bonow RH, et al. Long-Term Outcomes After Severe Acute Brain Injury Requiring Mechanical Ventilation: Recovery Trajectories Among Patients and Mental Health Symptoms of Their Surrogate Decision Makers. Neurocrit Care. 2025;42:896–910. doi: 10.1007/s12028-024-02164-2

4. Wendlandt B, Olm-Shipman C, Ceppe A, Hough CL, White DB, Cox CE, Carson SS. Surrogates of Patients With Severe Acute Brain Injury Experience Persistent Anxiety and Depression Over the 6 Months After ICU Admission. J Pain Symptom Manage. 2022;63:e633–e639. doi: 10.1016/j.jpainsymman.2022.02.336

5. Winstein CJ, Stein J, Arena R, Bates B, Cherney LR, Cramer SC, Deruyter F, Eng JJ, Fisher B, Harvey RL, et al. Guidelines for Adult Stroke Rehabilitation and Recovery: A Guideline for Healthcare Professionals From the American Heart Association/American Stroke Association. Stroke. 2016;47:e98–e169. doi: 10.1161/STR.0000000000000098

6. Miller EL, Murray L, Richards L, Zorowitz RD, Bakas T, Clark P, Billinger SA, Council AHACoCNatS. Comprehensive overview of nursing and interdisciplinary rehabilitation care of the stroke patient: a scientific statement from the American Heart Association. Stroke. 2010;41:2402–2448. doi: 10.1161/STR.0b013e3181e7512b

7. Bakas T, McCarthy MJ, Miller EL. Systematic Review of the Evidence for Stroke Family Caregiver and Dyad Interventions. Stroke. 2022;53:2093–2102. doi: 10.1161/STROKEAHA.121.034090

8. Kavalieratos D, Corbelli J, Zhang D, Dionne-Odom JN, Ernecoff NC, Hanmer J, Hoydich ZP, Ikejiani DZ, Klein-Fedyshin M, Zimmermann C, et al. Association Between Palliative Care and Patient and Caregiver Outcomes: A Systematic Review and Meta-analysis. JAMA. 2016;316:2104–2114. doi: 10.1001/jama.2016.16840

9. Greer JA, Jacobs JM, El-Jawahri A, Nipp RD, Gallagher ER, Pirl WF, Park ER, Muzikansky A, Jacobsen JC, Jackson VA, et al. Role of Patient Coping Strategies in Understanding the Effects of Early Palliative Care on Quality of Life and Mood. J Clin Oncol. 2018;36:53–60. doi: 10.1200/JCO.2017.73.7221

10. Chammas D, Moment A, Leff V, Buxton D, Rosa WE, Brenner K. Top Ten Tips Palliative Care Clinicians Should Know About Supporting Coping in Serious Illness. J Palliat Med. 2026;29:374–380. doi: 10.1089/jpm.2025.0223

11. National Academies of Sciences E, and Medicine, Health and Medicine Division, Board on Health Care Services, Committee on Integrating Social Needs Care into the Delivery of Health Care to Improve the Nation’s Health. Integrating Social Care into the Delivery of Health Care: Moving Upstream to Improve the Nation’s Health. National Academies Press (US). 2019. doi: PMID: 31940159.

12. Reichman M, Grunberg VA, Presciutti AM, Foster KT, Vranceanu AM, Creutzfeldt CJ. Peer-Delivered Interventions for Caregivers in the ICU with a Focus on Severe Acute Brain Injury: A Scoping Review. Neurocrit Care. 2024. doi: 10.1007/s12028-024-02115-x

13. Rutz Voumard R, Kiker WA, Dugger KM, Engelberg RA, Borasio GD, Curtis JR, Jox RJ, Creutzfeldt CJ. Adapting to a New Normal After Severe Acute Brain Injury: An Observational Cohort Using a Sequential Explanatory Design. Crit Care Med. 2021. doi: 10.1097/CCM.0000000000004947

14. Carroll C, Patterson M, Wood S, Booth A, Rick J, Balain S. A conceptual framework for implementation fidelity. Implement Sci. 2007;2:40. doi: 10.1186/1748-5908-2-40

15. Bellg AJ, Borrelli B, Resnick B, Hecht J, Minicucci DS, Ory M, Ogedegbe G, Orwig D, Ernst D, Czajkowski S, et al. Enhancing treatment fidelity in health behavior change studies: best practices and recommendations from the NIH Behavior Change Consortium. Health Psychol. 2004;23:443–451. doi: 10.1037/0278-6133.23.5.443

16. Miles MB, Huberman AM, Saldaña J. Qualitative data analysis : a methods sourcebook. Fourth edition. ed. Los Angeles: SAGE; 2020.

17. Ferrell B, Sun V, Hurria A, Cristea M, Raz DJ, Kim JY, Reckamp K, Williams AC, Borneman T, Uman G, et al. Interdisciplinary Palliative Care for Patients With Lung Cancer. J Pain Symptom Manage. 2015;50:758–767. doi: 10.1016/j.jpainsymman.2015.07.005

18. Li X, Hu S, Zhou Y, Ying X, Wu T. Impact of nurse-led palliative care on symptom management and life quality outcomes in elderly cancer patients: A retrospective study. Medicine (Baltimore). 2024;103:e39817. doi: 10.1097/MD.0000000000039817

19. Bekelman DB, Feser W, Morgan B, Welsh CH, Parsons EC, Paden G, Baron A, Hattler B, McBryde C, Cheng A, et al. Nurse and Social Worker Palliative Telecare Team and Quality of Life in Patients With COPD, Heart Failure, or Interstitial Lung Disease: The ADAPT Randomized Clinical Trial. JAMA. 2024;331:212–223. doi: 10.1001/jama.2023.24035

20. Kiker WA, Rutz Voumard R, Andrews LIB, Holloway RG, Brumback LC, Engelberg RA, Curtis JR, Creutzfeldt CJ. Assessment of Discordance Between Physicians and Family Members Regarding Prognosis in Patients With Severe Acute Brain Injury. JAMA Netw Open. 2021;4:e2128991. doi: 10.1001/jamanetworkopen.2021.28991

21. Sanders JJ, Curtis JR, Tulsky JA. Achieving Goal-Concordant Care: A Conceptual Model and Approach to Measuring Serious Illness Communication and Its Impact. J Palliat Med. 2018;21:S17–S27. doi: 10.1089/jpm.2017.0459

22. Halpern SD. Goal-Concordant Care - Searching for the Holy Grail. N Engl J Med. 2019;381:1603–1606. doi: 10.1056/NEJMp1908153

23. Bandura A. Social cognitive theory: an agentic perspective. Annu Rev Psychol. 2001;52:1–26. doi: 10.1146/annurev.psych.52.1.1

24. Segal O, Sher H, Aderka IM, Weinbach N. Does acceptance lead to change? Training in radical acceptance improves implementation of cognitive reappraisal. Behav Res Ther. 2023;164:104303. doi: 10.1016/j.brat.2023.104303

25. Shearin EN, Linehan MM. Dialectical behavior therapy for borderline personality disorder: theoretical and empirical foundations. Acta Psychiatr Scand Suppl. 1994;379:61–68. doi: 10.1111/j.1600-0447.1994.tb05820.x

